# Effect of in-hospital treatment with angiotensin-converting enzyme inhibitors and angiotensin II receptor blockers on mortality and complications in patients hospitalized for COVID-19: a large Spanish cohort study

**DOI:** 10.1101/2021.02.24.21252391

**Authors:** Emilia Roy-Vallejo, Aquilino Sánchez Purificación, José David Torres Peña, Beatriz Sánchez Moreno, Francisco Arnalich, María José García Blanco, José López Miranda, Juan Luis Romero Cabrera, Carmen Rosario Herrero Gil, J Bascunana, Manuel Rubio-Rivas, Sara Pintos Otero, Verónica Martínez Sempere, Jesús Ballano Rodríguez-Solís, Ricardo Gil Sánchez, Jairo Luque del Pino, Amara González Noya, MS Navas-Alcántara, Begoña Cortés Rodríguez, José N. Alcalá, Ana Suárez-Lombraña, Jorge Andrés Soler, Ricardo Gómez Huelgas, José Manuel Casas Rojo, Jesús Millán Núñez-Cortés, for the SEMI-COVID-19 Network^

## Abstract

**Background:** The use of ACEI (Angiotensin-Converting Enzyme Inhibitor) and ARB (Angiotensin II Receptor Blocker) in COVID-19 remains controversial. Our main aim was to describe the effect of ACEI/ARB treatment during COVID-19 hospitalization on mortality and complications.

**Methods:** Retrospective, observational, multicenter study, part of the SEMI-COVID-19 Registry, comparing patients with COVID-19 treated with ACEI/ARB during hospitalization to those not treated. The primary endpoint was incidence of the composite outcome of prognosis (IMV [Invasive Mechanical Ventilation], NIMV [Non-Invasive Mechanical Ventilation], ICU admission [Intensive Care Unit], and/or all-cause mortality). The secondary endpoint was incidence of MACE (Major Adverse Cardiovascular Events). We evaluated both outcomes in patients whose treatment with ACEI/ARB continued or was withdrawn during hospitalization.

**Results:** Between February and June 2020, 11,205 patients were included, with mean age 67 years (SD=16.3) and 43.1% female; 2,162 patients received ACEI/ARB treatment. ACEI/ARB treatment showed a protective effect on all-cause mortality (*p*<.0001). In hypertensive patients it was also protective in terms of IMV, ICU admission, and the composite outcome of prognosis (*p*<.0001 for all). No differences were found in incidence of MACE. Patients previously treated with ACEI/ARB who continued treatment during hospitalization had a lower incidence of the composite outcome of prognosis than those whose treatment was withdrawn (RR 0.67, 95%CI 0.63-0.76). ARB had a more beneficial effect on survival than ACEI (HR 0.77, 95%CI 0.62-0.96).

**Conclusion:** ACEI/ARB treatment during COVID-19 hospitalization had a protective effect on mortality. The benefits were greater in hypertensive patients, those who continued treatment during hospitalization, and those taking ARB.

**Summary:** Treatment with ACEI/ARB during COVID-19 hospitalization showed a beneficial effect on mortality in the general population. The benefit was greater in hypertensive patients, in those who maintained treatment during hospitalization and those taking ARB.

## Introduction

It has been suggested that the renin-angiotensin-aldosterone system (RAAS) has a relevant role in the pathogenesis of COVID-19, caused by SARS-CoV-2. The virus can infect host cells through interaction with angiotensin-converting enzyme 2 (ACE-2) in the respiratory epithelium [1]. Therefore, there has been concern about risk related to the use of drugs that act on the RAAS in the context of the COVID-19.

This concern is even more relevant given that a large part of patients with COVID-19 are hypertensive. Different case series have shown that hypertension, diabetes, and cardiovascular disease—pathologies in which these drugs are usually prescribed—are more common in COVID-19 patients with severe disease than in those with mild or moderate disease [2]. Several publications provide data on whether the use of ACEI or ARB are harmful in the context of SARS-CoV-2 infection, though they are all observational studies, and the effect of these drugs is not clear [1,3]. To date, until more evidence emerges, the main scientific societies [4,5] advise against the interruption of these drugs.

SARS-CoV-2 has a high morbidity and mortality rate. As of November 28, 2020, nearly 62 million (61,967,977) cases of COVID-19 had been diagnosed and over 1,448,411 people had died [6]. Therefore, the analysis of large datasets is essential to establish patient profiles so that the currently available therapeutic tools can be used and it can be ascertained whether treatment before or during the infection can alter the course of the disease.

Until now, the effect of ACEI and ARB use in patients with SARS-CoV-2 infection during hospitalization on mortality and other prognostic outcomes had yet to be evaluated in a Spanish cohort.

The main objective of this work was to evaluate how treatment with ACEI/ARB in hospitalized SARS-CoV-2 patients in Spain alters prognosis. As a secondary objective, we analyzed whether treatment with these medications during hospitalization had an effect on the incidence of major adverse cardiovascular events (MACE).

## Material and methods

### Study design

Observational, retrospective, multicenter study that is part of the SEMI-COVID-19 Registry. This registry is an open initiative of the Spanish Society of Internal Medicine (SEMI) with 150 participant hospitals. The registry enrolls consecutive patients with a positive SARS-CoV-2 reverse transcription polymerase chain reaction (RT-PCR) of a nasopharyngeal swab admitted to participating hospitals in Spain since February 2020. This work includes completed data compiled as of June 4, 2020.

The inclusion criteria for the study were the following: age > 18 years, positive RT-PCR for SARS-CoV-2, admission to any participating hospital, and availability of age, sex, race, and onset of symptoms. We excluded patients who remained hospitalized as of June 4, 2020, those who did not have data available on treatment with ACEI/ARB before and during hospitalization, and those without the date of the first positive RT-PCR recorded (Figure 1).

**Figure 1.**
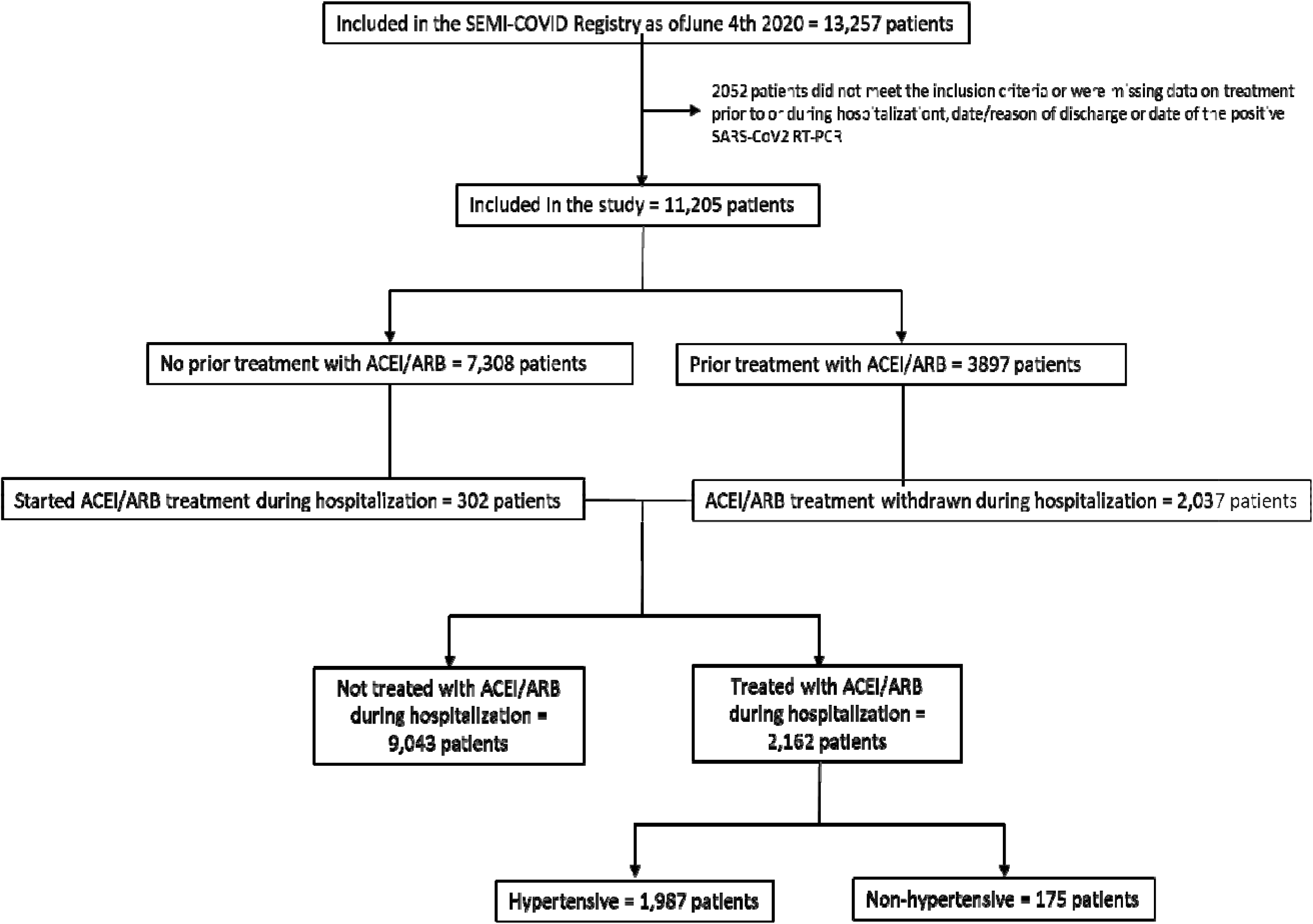
Patient inclusion flowchart.

Patients were classified in the ACEI/ARB group if they received at least one dose of any ACEI/ARB during hospitalization. Previous ACEI/ARB use was determined based on whether it was recorded in the last entry in the medical chart. Each patient’s management and treatment were the responsibility of the attending physicians.

### Data collection

All data were collected from the medical charts and included in the registry’s encrypted online database. An independent external agency monitored and reviewed the information for inconsistencies. Full details on the SEMI-COVID-19 Registry have been described previously [7].

### Main outcomes

In order to evaluate the study’s primary objective, we created a composite outcome of prognosis that included the need for invasive mechanical ventilation (IMV), non-invasive mechanical ventilation (NIMV), intensive care unit (ICU) admission, and/or all-cause mortality.

As secondary objectives, we assessed the effect of ACEI/ARB treatment on cardiovascular risk in the context of COVID-19. Then, we established the composite outcome of major adverse cardiovascular events (MACE), which included incidence of myocardial infarction (MI), heart failure (HF), stroke, and/or any arrhythmia (atrial or ventricular). Cardiovascular mortality was not included because these data were not available in the Registry.

### Statistical analysis

Quantitative variables were expressed as mean and standard deviation (SD) or median and interquartile range (IQR) if they did not follow a Gaussian distribution. Categorical variables were expressed as frequencies and percentages. Baseline characteristics were compared between the ACEI/ARB and non-ACEI/ARB groups using either Student’s t-test or the Wilcoxon test for continuous variables or the chi-square test for qualitative variables.

We compared the incidence of both the aforementioned composite outcomes and of each individual event during hospitalization between the ACEI/ARB and non-ACEI/ARB groups using logistic regression models and estimating adjusted relative risks using the marginal standardization method. On each regression analysis, several predictors were considered to be possible modifying and/or confounding factors: age, sex, race, smoking and alcohol use, hypertension, dyslipidemia, diabetes mellitus, obesity, chronic kidney disease, chronic heart failure, prior treatment with ACEI/ARB, Charlson Comorbidity Index, and in-hospital treatment with tocilizumab or corticosteroids. For the composite outcomes, we selected the most parsimonious models which did not result in a clinically significant change (< 5%) on the odds ratio in comparison to the maximal reference model. We used this more restrictive threshold because there were many models to compare, given the number of predictors evaluated.

Using logistic regression models, we analyzed whether the effect of ACEI/ARB use was sustained in patients whose treatment was withdrawn compared to those whose ACEI/ARB treatment was continued. On this analysis, we considered the same composite outcomes of prognosis and MACE.

Lastly, we performed a survival analysis to evaluate the effect of ACEI/ARB use and of continuing/withdrawing this treatment during hospitalization, comparing Kaplan-Meier curves with the logrank test. In addition, Cox proportional-hazards models allowed for including additional covariates in a similar manner to the logistic regression analysis described above. These results were expressed as hazard ratios after confirming that the proportional hazard assumption was met.

All statistical analyses were performed using Stata software (version 15.0, Stata Corp, College Station, TX, USA). A two-tailed *p* value of 5% was established as the threshold of statistical significance.

### Ethics

The SEMI-COVID-19 Registry was evaluated and approved by the Provincial Research Ethics Committee of Málaga. Given the state of emergency declared during the pandemic, it was only mandatory for patients to provide verbal consent. This manuscript was written following the recommendations of the Strengthening the Reporting of Observational studies in Epidemiology (STROBE) Statement.

## Results

### Study population

A total of 11,205 patients were included in the study (Fig. 1). Their baseline characteristics are shown in Table 1, treatment and analytical data are shown in Supplementary table 1. The mean age of patients was 67 years (SD=16.3), 43.1% were female, 89.3% were Caucasian, and the mean age-adjusted Charlson Comorbidity Index was 3.6 points (SD=2.7). During hospitalization, 2,162 (19.3%) participants were treated with ACEI/ARB. Subjects in the ACEI/ARB group were older (72.5 vs 65.7 years, *p*<0.0001), more frequently male (59.1% vs 56.3%, *p*=0.018) and Caucasian (93.8% vs 88.2%, *p*<0.001) than those in the non-ACEI/ARB group. Furthermore, they had more comorbidities (age-adjusted Charlson Comorbidity Index of 4.4 points (SD=2.5) vs 3.4 points (SD=2.7), *p*<0.001), with hypertension being the most prevalent (92.1% vs 39.7%, *p*<0.001). Prior to hospitalization, 41.5% of patients were treated with ACEI and 47.6% with ARB in the ACEI/ARB group, versus 16.9% and 19%, respectively in the non-ACEI/ARB group. The main reason for ACEI/ARB treatment (92% of the patients) was hypertension, while 8% were taking these drugs for other diseases.

**Table 1.** Demographic and clinical data.

|  | Total population<br>(n = 11,205) | Non-ACEI/ARB group<br>(n = 9,043) | ACEI/ARB group<br>(n = 2,162) | p value |
| --- | --- | --- | --- | --- |
| Age (years): mean (SD) | 67.0 (16.3) | 65.7 (16.8) | 72.5 (12.5) | <0.0001 |
| Female sex (%) | 4,827/11,190 (43.1%) | 3,944/9,030 (43.7%) | 883/2,160 (40.9%) | 0.018 |
| Race/Ethnicity (%) |  |  |  | <0.001* |
| • Caucasian | 9,831/11,014 (89.3%) | 7836/8,886 (88.2%) | 1,995/2,128 (93.8%) |  |
| • African | 45/11,014 (0.4%) | 36/8,886 (0.4%) | 9/2,128 (0.4%) |  |
| • Latin American | 990/11,014 (9.0%) | 892/8,886 (10.0%) | 98/2,128 (4.6%) |  |
| • Asian | 50/11,014 (0.5%) | 43/8,886 (0.5) | 7/2,128 (0.3%) |  |
| • Other | 98/11,014 (0.9%) | 79/8,886 (0.9%) | 19/2,128 (0.9%) |  |
| Smoking (%) |  |  |  | <0.001* |
| • Non-smoker | 7,438/10,699 (69.5%) | 6,125/8,617 (71.1%) | 1,313/2,082 (63.1%) |  |
| • Former smoker | 2,686/10,699 (25.0%) | 2,030/8,617 (23.6%) | 656/2,082 (31.5%) |  |
| • Active smoker | 575/10,699 (5.4%) | 462/8,617 (5.4%) | 113/2,082 (15.4%) |  |
| Alcohol use disorder (%) | 516/10,887 (4.7%) | 387/8,780 (4.4%) | 129/2,107 (6.1%) | 0.001 |
| Comorbidities (%) |  |  |  |  |
| • Hypertension | 5,576/11,190 (49.8%) | 3,589/9,033 (39.7%) | 1,987/2,157 (92.1%) | <0.001 |
| • Dyslipidemia | 4,415/11,189 (39.5%) | 3,231/9,029 (35.8%) | 1,184/2,160 (54.8%) | <0.001 |
| • Diabetes mellitus | 2,095/11,178 (18.7%) | 1,464/9,022 (16.2%) | 631/2,156 (29.3%) | <0.001 |
| • Obesity | 2,186/10,212 (21.4%) | 1,618/ 8,242 (19.6%) | 568/1,970 (28.8%) | <0.001 |
| • Heart failure | 811/11,186 (7.3%) | 594/ 9,030 (6.6%) | 217/2,156 (10.1%) | <0.001 |
| • Ischemic heart disease | 880/11,193 (7.9%) | 591/ 9,032 (6.5 %) | 289/2,161 (13.4%) | <0.001 |
| • Cerebrovascular disease | 797/11,173 (7.1%) | 598/9,018 (6.6%) | 199/2,155 (9.2%) | <0.001 |
| • Peripheral artery disease | 523/11,183 (4.7%) | 387/9,026 (4.3%) | 136/2,157 (6.3%) | <0.001 |
| • Chronic kidney disease | 665/11,183 (5.9%) | 491/ 9,026 (5.4%) | 174/2,157 (8.1%) | <0.001 |
| - Age-adjusted Charlson Comorbidity Index: points (SD) | 3.6 (2.7) | 3.4 (2.7) | 4.4 (2.47) | <0.0001 |
| Previous treatment (%) |  |  |  |  |
| • ACEI | 1,890/11,205 (16.9%) | 993/ 9,043 (11.0%) | 897/2,162 (41.5%) | <0.001 |
| • ARB | 2,133/11,205 (19.0%) | 1,103/9,043 (12.2 %) | 1,030/2,162 (47.6%) | <0.001 |

SD: Standard Deviation, IQR: Interquartile Range, ACEI: Angiotensin-Converting Enzyme Inhibitor, ARB: Angiotensin II Receptor Blocker.
\*The *p* value refers to Caucasian and Latin American patients and all categories of smoking.

### Outcomes of prognosis

At least one of the events included in the composite outcome of prognosis (IMV, NIMV, ICU admission, or death) occurred in 569 patients (27.0%) in the ACEI/ARB group and in 2,443 (27.6%) in the non-ACEI/ARB group (*p*=0.6). The results of the univariate and multivariate analyses are shown in Table 2.

**Table 2.** Univariate and multivariate analysis of prognosis outcomes by treatment group

| Outcome | Non-ACEI/ARB group | ACEI/ARB group | Univariate |  | Multivariate |  |
| --- | --- | --- | --- | --- | --- | --- |
|  |  |  | RR (95%CI) | p value | RR (95%CI) | p value |
| Composite variable of prognosis: IMV, NIMV, ICU admission, or death* | 2,443/8,854 (27.6%) | 569/2,107 (27.0%) | 0.98 (0.91-1.06) | 0.6064 | Normotensive: 1.12 (0.90-1.38)<br>Hypertensive: 0.68 (0.62-0.75) | 0.3085<br><0.0001 |
| IMV* | 622/9,024 (6.9%) | 115/2,157 (5.3%) | 0.77 (0.64-0.94) | 0.0100 | Normotensive: 2.07 (1.42-3.02)<br>Hypertensive: 0.50 (0.39-0.64) | 0.0002<br><0.0001 |
| NIMV | 396/9,026 (4.4%) | 130/2,156 (6.0%) | 1.37 (1.13-1.67) | 0.0015 | 1.13 (0.90-1.43) | 0.2908 |
| ICU admission* | 767/9,035 (8.5%) | 148/2,161 (6.8%) | 0.81 (0.68-0.96) | 0.0140 | Normotensive: 1.76 (1.23-2.52)<br>Hypertensive: 0.57 (0.46-0.71) | 0.0019<br><0.0001 |
| Death | 1,897/8,853 (21.4%) | 436/2,105 (20.7%) | 0.97 (0.88-1.06) | 0.4897 | 0.65 (0.59-0.72) | <0.0001 |

\*A significant relationship with hypertension was found on the multivariate analysis
RR: Relative Risk, 95%CI: 95% Confidence Interval, ACEI: Angiotensin-Converting Enzyme Inhibitor, ARB:
Angiotensin II Receptor Blocker, IMV: Invasive Mechanical Ventilation, NIMV: Non-Invasive Mechanical
Ventilation, ICU: Intensive Care Unit.

The multivariate analysis revealed that patients treated with ACEI/ARB during hospitalization had lower mortality risk (RR 0.65, 95%CI 0.59-0.72) but had no significant effect on the probability of needing NIMV (RR 1.13, 95%CI 0.9-1.43).

We found different effects in hypertensive and non-hypertensive patients. In the hypertensive subgroup, treatment with ACEI/ARB during hospitalization had a protective effect in terms of the composite variable of prognosis (RR 0.68, 95%CI 0.62-0.75), IMV (RR 0.50, 95%CI 0.39-0.64), and ICU admission (RR 0.57, 95%CI 0.46-0.71). In the normotensive subgroup, it showed a neutral effect on the composite variable of prognosis (RR 1.12, 95%CI 0.90-1.38) and was associated with a higher risk of IMV (RR 2.07, 95%CI 1.42-3.02) and ICU admission (RR 1.76, 95%CI 1.23-2.52). Neither NIMV nor all-cause mortality had a significant relationship with hypertension or any other predictors (reduced regression model in supplementary table 2).

On the survival analysis, significant differences between the treatment groups were found when patients were classified by hypertensive status (Fig. 2).

**Figure 2.**
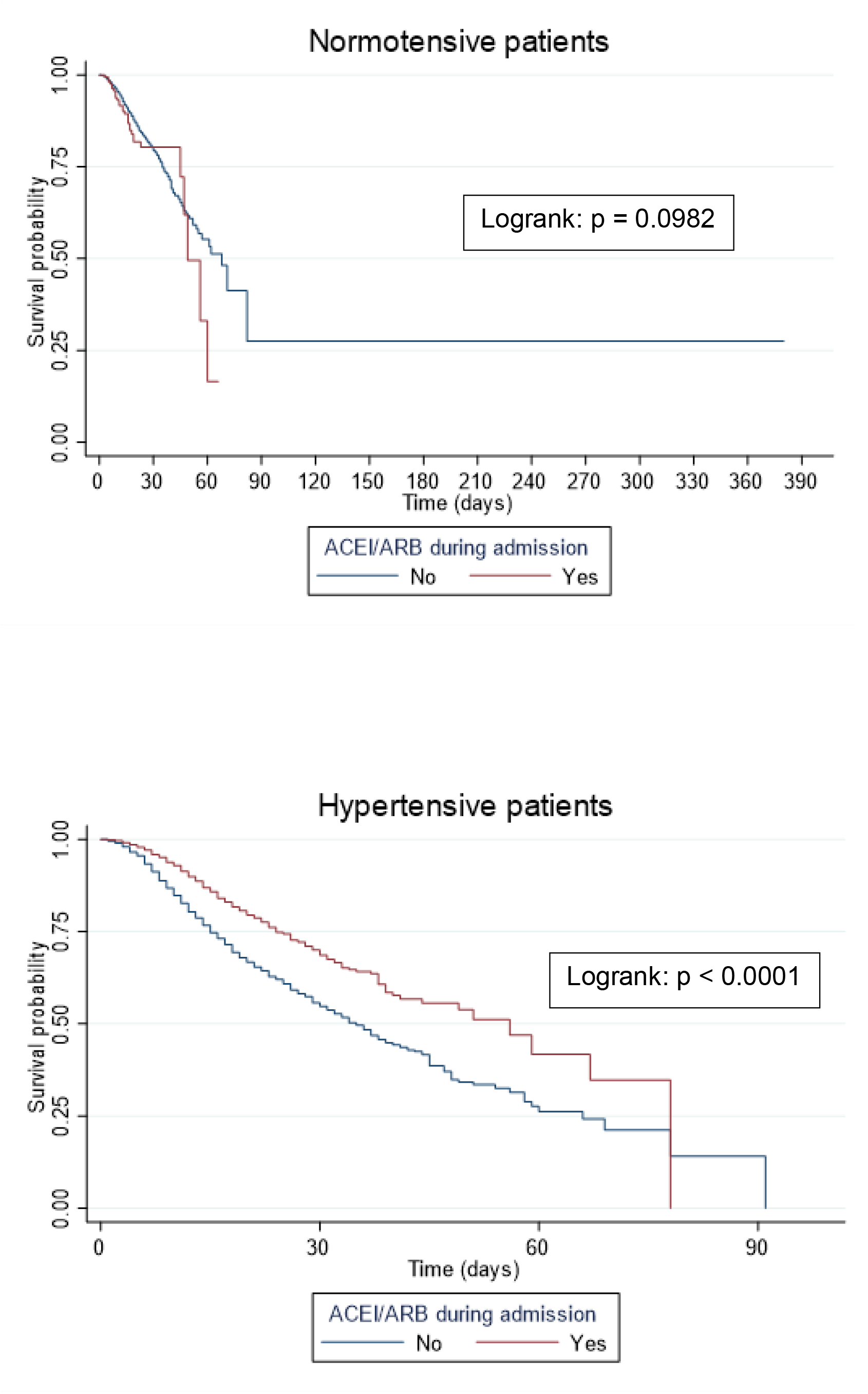
Effect of treatment with ACEI/ARB during hospitalization: Kaplan-Meier curves adjusted for hypertension.

We also performed a Cox regression and since it did not meet the proportionality assumption, we calculated the HR for different periods of time starting with onset of symptoms. The protective effect of ACEI/ARB treatment progressively decreased over time (Table 3).

**Table 3.**
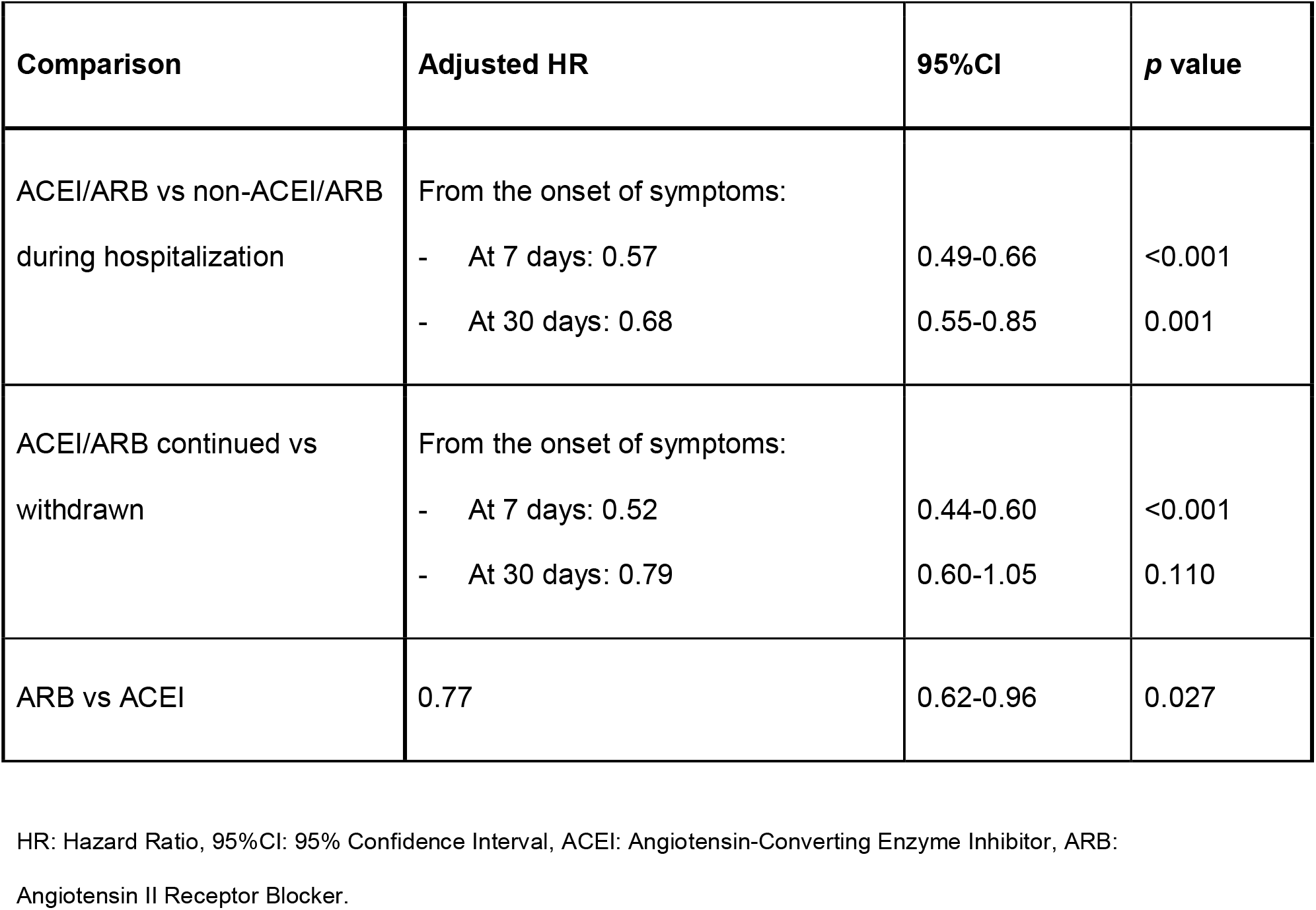
Survival analysis: Hazard ratios (HR) estimated via Cox regression.

### Major Adverse Cardiovascular Events

In the ACEI/ARB group, 12% of patients had a MACE, significantly higher than the 9.2% found in the non-ACEI/ARB group (*p*=0.001). A description of the MACE by group and analysis are shown in Table 4.

**Table 4.**
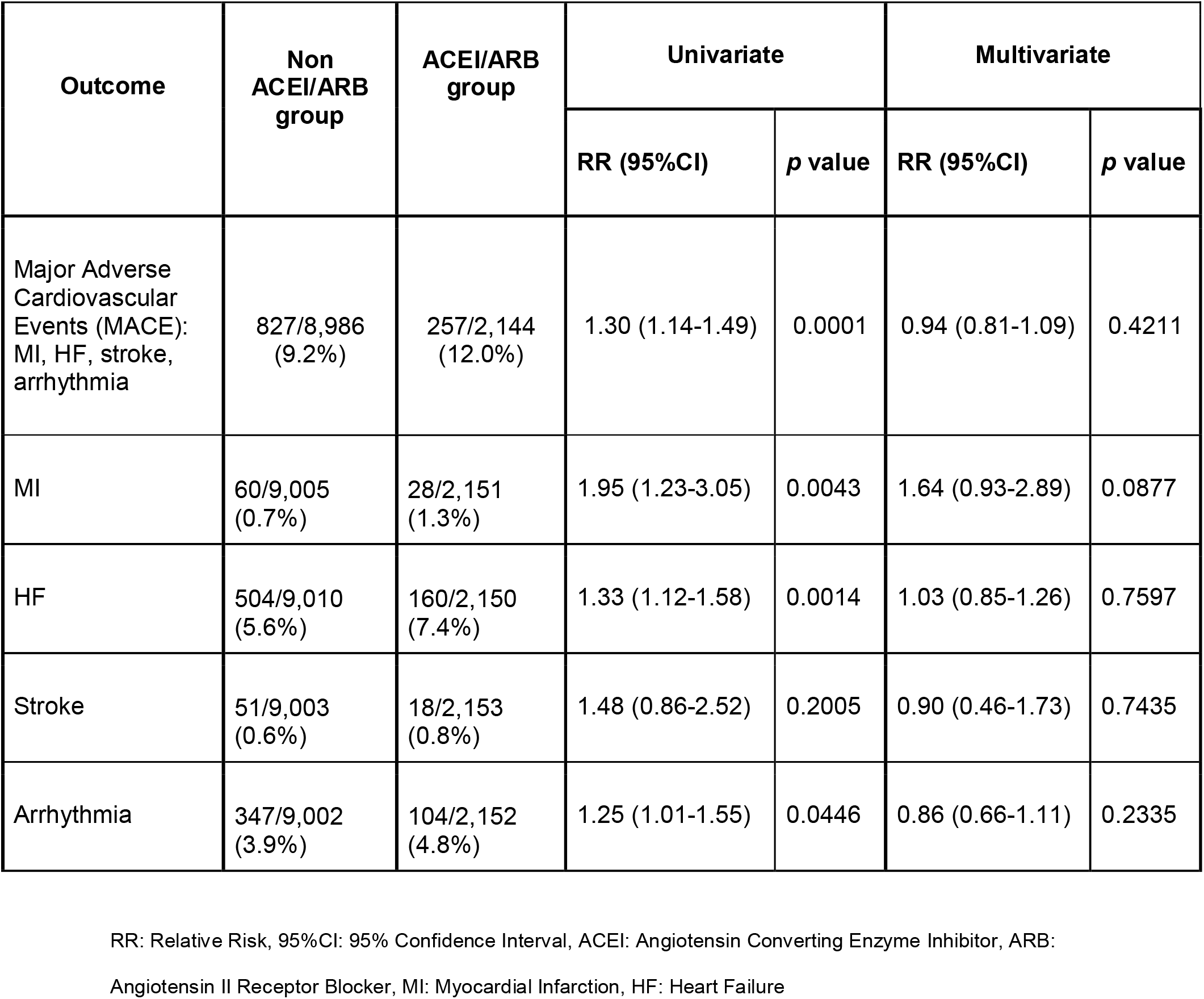
Univariate and multivariate analysis of MACE outcomes by treatment group.

| Outcome | Non ACEI/ARB group | ACEI/ARB group | Univariate |  | Multivariate |  |
| --- | --- | --- | --- | --- | --- | --- |
|  |  |  | RR (95%CI) | *p* value | RR (95%CI) | *p* value |
| Major Adverse Cardiovascular Events (MACE): MI, HF, stroke, arrhythmia | 827/8,986 (9.2%) | 257/2,144 (12.0%) | 1.30 (1.14-1.49) | 0.0001 | 0.94 (0.81-1.09) | 0.4211 |
| MI | 60/9,005 (0.7%) | 28/2,151 (1.3%) | 1.95 (1.23-3.05) | 0.0043 | 1.64 (0.93-2.89) | 0.0877 |
| HF | 504/9,010 (5.6%) | 160/2,150 (7.4%) | 1.33 (1.12-1.58) | 0.0014 | 1.03 (0.85-1.26) | 0.7597 |
| Stroke | 51/9,003 (0.6%) | 18/2,153 (0.8%) | 1.48 (0.86-2.52) | 0.2005 | 0.90 (0.46-1.73) | 0.7435 |
| Arrhythmia | 347/9,002 (3.9%) | 104/2,152 (4.8%) | 1.25 (1.01-1.55) | 0.0446 | 0.86 (0.66-1.11) | 0.2335 |

RR: Relative Risk, 95%CI: 95% Confidence Interval, ACEI: Angiotensin Converting Enzyme Inhibitor, ARB:
Angiotensin II Receptor Blocker, MI: Myocardial Infarction, HF: Heart Failure

After adjusting for confounding variables, treatment with ACEI/ARB during hospitalization was found to be neither protective nor harmful regarding the composite outcome of MACE or any of its components (reduced regression model in supplementary table 2).

### ACEI/ARB continuation versus withdrawal during hospitalization

A total of 3,897 patients were receiving ACEI and/or ARB prior to hospitalization; treatment was continued in 1,860 of them. The ACEI/ARB group had a lower probability of the composite variable of prognosis (RR 0.66, 95%CI 0.6-0.72) and MACE (RR 0.75, 95%CI 0.64-0.88) compared to the non-ACEI/ARB group. This effect was sustained on the multivariate analysis for the composite variable of prognosis (RR 0.67, 95%CI 0.63-0.76) but not for MACE (RR 0.86, 95%CI 0.73-1.01).

There was also a significant difference on the survival analysis (Fig. 3) and continuing ACEI/ARB treatment led to a lower risk of death on the Cox regression, but this protective effect also diminished progressively over time (Table 3).

**Figure 3.**
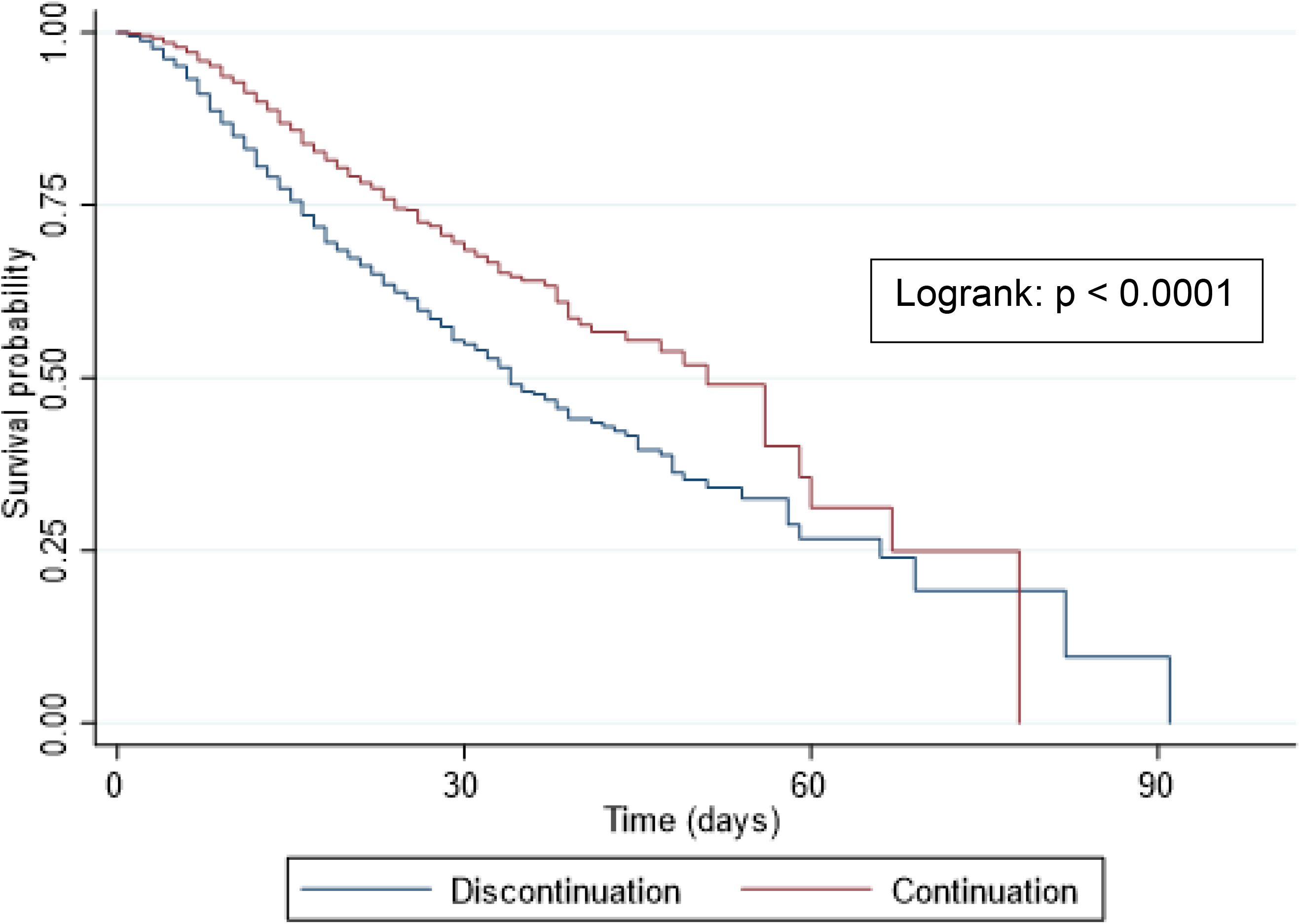
Effect of discontinuation vs continuation of ACEI/ARB treatment during hospitalization: Kaplan-Meier curves.

### Comparison between ACEI and ARB

When comparing the effects of ACEI versus ARB during hospitalization, we found only differences on survival with a more favorable effect with ARB after adjusting for confounding factors (Table 3).

## Discussion

In this series, we analyzed the effects of ACEI/ARB in a large number of patients admitted to Spanish hospitals for COVID-19 between February and June 2020. The most relevant finding of this study is that ACEI/ARB treatment during hospitalization had a protective effect, reducing mortality by 33%. No significant differences were found between the ACEI/ARB and non-ACEI/ARB groups on the composite outcome of prognosis; however, on the univariate analysis, there was a significant reduction in the relative risk on some of its components: 19% on ICU admission and 23% on need for IMV. A lack of significant difference was only found in the need for NIMV. Notwithstanding, it is important to note that during the period studied, Spanish hospitals were overrun and in many cases, ICUs had to triple their capacity [8,9]. Therefore, NIMV was reserved for patients with a poorer prognosis and a lower chance of survival.

It is especially relevant to highlight the benefits observed with ACEI/ARB in hypertensive patients with COVID-19. Indeed, 92.1% of those who received these drugs during hospitalization were hypertensive. Adjusted results showed a relative risk reduction (RRR) of 32% on the composite variable of prognosis in these patients. There was also a significant benefit observed on the different prognostic variables: 50% RRR for IMV, 43% for ICU admission, and 35% for all-cause mortality. These are better results than those obtained with other treatments used in hospitalized patients with COVID-19 [10].

The results obtained with ACEI/ARB are even more noteworthy if we consider that the group of patients who received these drugs had a higher mean age, greater comorbidity and a higher prevalence of cardiovascular diseases and vascular risk factors (Table 1). It has been published in this Registry [11] and in other series [12] that age, hypertension, and previous cardiovascular disease are all factors associated with a worse prognosis in patients with SARS-CoV-2.

This study also includes a small proportion of patients (8%; 170 patients) who received ACEI/ARB for diseases other than hypertension, including heart failure, coronary heart disease, and chronic kidney disease. However, we were not able to determine the cause of treatment in a significant number of patients. These diseases put patients at a high risk of complications and mortality from any hospital admission, not just for COVID-19. In most, it does not seem essential to maintain these treatments during hospitalization and indeed, if they are maintained, they might cause adverse effects such as hypotension, acute renal failure, or electrolyte imbalance. This likely explains the worse prognosis found in this subgroup and could be an explanation for why the results observed in the entire study population are not as unequivocal as in the hypertensive population. For this reason, some authors have excluded patients with severe organ dysfunction and patients with hypotension from their series [13].

Our results are consistent with other published series of patients hospitalized with COVID-19 who were treated with ACEI/ARB. Reynolds et al. [1] analyzed patients treated with ACEI/ARB and did not find a greater likelihood of a positive SARS-CoV-2 test or greater risk of severe COVID-19. Mancia et al. [3] showed no association between the use of ACEI/ARB and risk of contracting COVID-19 or a severe course of the disease. Like our work, both studies defined the need for mechanical ventilation or death as severe illness and the treated group also had a worse clinical profile and higher prevalence of cardiovascular diseases.

It should be noted that the survival curves show a clear benefit to maintaining previous treatment with ACEI/ARB in hypertensive patients during hospitalization (Figure 3), though this benefit disappears over the long term (>30 days). However, it is likely that patients with longer hospital stays had a poor prognosis due to complications, regardless of the treatment received. In studies analyzing 28-day mortality, Zhou et al. [13] and Zhang et al. [14] also found that in-hospital use of ACEI/ARB was associated with a significantly lower risk of mortality, notably among patients with hypertension.

In this series, some treatments could have been withdrawn due to complications, but in most cases ACEI/ARB were suspended upon admission because of concerns about their safety. Our results reinforce the idea that ACEI/ARB are beneficial drugs in hypertensive patients with COVID-19: their withdrawal during hospitalization leads to a greater risk for complications and mortality.

No benefits were observed in terms of a reduction in MACE by maintaining treatment with ACEI/ARB during hospitalization for COVID-19 (Table 3). Though the beneficial effects of ACEI/ARB have been widely demonstrated for the cardiovascular events included in MACE [15–17], a hospitalization is not enough time to evaluate the cardiovascular benefit of treatments. The higher mean age and higher prevalence of cardiovascular risk factors and previous cardiovascular diseases in the ACEI/ARB group may explain the greater incidence of cardiovascular events observed. This result seems to indicate that the benefit achieved with ACEI/ARB during COVID-19 hospitalization is in fact not cardiovascular. Some studies suggest that these drugs lead to a lesser inflammatory response during acute lung injury due to blockage of the renin-angiotensin-aldosterone system (RAAS), especially with ARB [18,19].

Though no differences were noted between ACEI and ARB on the main composite outcomes, a significant effect on survival was found in the group that received ARB (23% reduction in mortality). A recently published article found a similar result with ARB treatment [11]. ACEI and ARB act on different points of the RAAS and their effects are not the same. Although current data is limited, inconclusive, and based on animal models, there is some evidence that ARB may have a beneficial effect in reducing angiotensin II-induced alveolar permeability, an effect not observed with ACEI [18,20].

The novelty of this study is that we have assessed not just mortality but also severe respiratory complications as the primary outcomes. The need for mechanical ventilation or ICU admission were the worst complications patients had during COVID-19 hospitalization.

Our study has several strengths. First, it is one of the largest series of patients with COVID-19 published to date. As a nationwide, multicenter study, it quite accurately reflects the reality in Spain during the first months of the pandemic. Second, on the data analysis, we included hypertensive patients as well as all who received ACEI/ARB during hospitalization for COVID-19. In addition to mortality, major respiratory and cardiovascular complications recorded during hospitalization were included as one of the main objectives of this study and we analyzed the effect that discontinuation of these drugs had on these complications. All of this allows for a better evaluation of the effect of ACEI/ARB on patients’ prognosis.

This study also has several limitations. First, its retrospective and observational nature. Second, the decision to maintain or withdraw treatment during hospitalization depended on each attending physician’s judgment, leading to selection bias. Third, we do not know which ACEI or ARB drugs were used, at what doses, and for how long. Lastly, this series mainly comprises Caucasian patients, so our results cannot be extrapolated to other populations.

In conclusion, our results show that maintaining treatment with ACEI/ARB in patients hospitalized for COVID-19 led to lower rates of death and respiratory complications, especially in hypertensive patients.

Prospective and randomized controlled trials are needed to confirm these results. This work also points to an exciting field of research to be explored further: analysis of the molecular mechanisms that underlie the protective effect of ACEI/ARB against SARS-CoV-2.

## Supporting information

Supplementary #x002B; STROBE

## Data Availability

All data will be available under request.

## Acknowledgements

We gratefully acknowledge all the investigators who participate in the SEMI-COVID-19 Registry. We also thank the SEMI-COVID-19 Registry Coordinating Center, S&H Medical Science Service, for their quality control data, logistic and administrative support.

