## Supplementary #x002B; STROBE for "Effect of in-hospital treatment with angiotensin-converting enzyme inhibitors and angiotensin II receptor blockers on mortality and complications in patients hospitalized for COVID-19: a large Spanish cohort study"

**SUPPLEMENTARY MATERIAL**:

Supplementary table 1. Laboratory and treatment data according to ACEI/ARB treatment group

| | Total population (n = 11,205) | Non-ACEI/ARB group (n = 9,043) | ACEI/ARB group (n = 2,162) | *p* value |
|---|---|---|---|---|
| Laboratory data | | | | |
| - Leukocytes (x$10^3$/mm$^3$): mean (SD) | 7,308.4 (5,469.8) | 7,263.9 (5,195.9) | 7,494.7 (6,490.9) | .0793 |
| - Lymphocytes (x$10^3$/mm$^3$): median (IQR) | 940 (600) | 930 (600) | 950 (600) | .5841 |
| - C-reactive protein (mg/dL): mean (SD) | 86.0 (87.5) | 86.3 (88.7) | 85.1 (82.3) | .5864 |
| - Procalcitonin (ng/mL): median (IQR) | 0.1 (0.2) | 0.1 (0.2) | 0.1 (0.1) | .5585 |
| - Creatinine (mg/dL): median (IQR) | 0.9 (0.4) | 0.89 (0.4) | 1 (0.5) | <.0001 |
| - Lactate dehydrogenase (U/L): mean (SD) | 358.7 (209.1) | 361.1 (220.0) | 348.6 (156.4) | .0042 |
| - Ferritin (mcg/L): median (IQR) | 612.7 (928) | 625 (949.5) | 569 (870) | .0941 |
| - Interleukin 6 (pg/mL): median (IQR) | 29.8 (53) | 28 (50.07) | 38.4 (57.6) | .0049 |
| - D-dimer (ng/mL): median (IQR) | 624 (797) | 610 (787.5) | 682 (879) | <.0001 |
| Treatment during hospitalization | | | | |
| - Corticosteroids (%) | 3,903/11,205 (34.8%) | 3,074/9,043 (34.0%) | 829/2,162 (38.3%) | <.001 |

| | | | | |
|---|---|---|---|---|
| - Total corticosteroid dose (mg prednisone): mean (SD) | 646.07 (564.8) | 639.6 (10.7) | 669.8 (20.5) | .1924 |
| - Tocilizumab (%) | 1,007/11,205 (9%) | 796/9,043 (8.8%) | 211/2,162 (9.8%) | .162 |

SD: Standard Deviation, IQR: Interquartile Range, ACEI: Angiotensin-Converting Enzyme Inhibitor, ARB: Angiotensin II Receptor Blocker.

Supplementary table 2. Reduced logistic regression models.

| **Dependent variable** | **Independent variables** | **Coefficients** | **95%CI** | ***p* value** | **OR** | **95%CI** | ***p* value** |
|---|---|---|---|---|---|---|---|
| **Composite outcome of prognosis** | **ACEI/ARB during admission** | 0.202 | -0.163 to 0.567 | .279 | 1.223 | 0.849 to 1.762 | .279 |
| | Hypertension | 0.178 | 0.045 to 0.312 | .009 | 1.195 | 1.046 to 1.366 | .009 |
| | **ACEI/ARB* Hypertension** | -0.803 | -1.189 to -0.417 | <.001 | 0.448 | 0.305 to 0.659 | <.001 |
| | Age | 0.050 | 0.047 to 0.054 | <.001 | 1.052 | 1.048 to 1.056 | <.001 |
| | Previous ACEI/ARB treatment | 0.142 | 0.010 to 0.273 | .035 | 1.152 | 1.010 to 1.314 | .035 |
| | Charlson Comorbidity Index | 0.178 | 0.153 to 0.203 | <.001 | 1.194 | 1.165 to 1.225 | <.001 |
| | Tocilizumab | 1.798 | 1.648 to 1.949 | <.001 | 6.040 | 5.197 to 7.020 | <.001 |
| | *Constant* | -5.024 | -5.295 to -4.753 | <.001 | 0.007 | 0.005 to 0.009 | <.001 |
| **Major Adverse Cardiovascular** | **ACEI/ARB during hospitalization** | -0.153 | -0.315 to 0.009 | .064 | 0.858 | 0.729 to 1.009 | .064 |

| | | | | | | | |
|---|---|---|---|---|---|---|---|
| **Events** | Age | 0.050 | 0.045 to 0.056 | <.001 | 1.051 | 1.046 to 1.058 | <.001 |
| | Smoking | | | | | | |
| | - Former smoker | 0.301 | 0.157 to 0.446 | <.001 | 1.352 | 1.170 to 1.562 | <.001 |
| | - Active smoker | 0.505 | 0.212 to 0.798 | .001 | 1.657 | 1.236 to 2.221 | .001 |
| | Hypertension | 0.572 | 0.411 to 0.732 | <.001 | 1.771 | 1.509 to 2.080 | <.001 |
| | Tocilizumab | 0.836 | 0.631 to 1.040 | <.001 | 2.307 | 1.880 to 2.830 | <.001 |
| | *Constant* | -6.355 | -6.787 to -5.924 | <.001 | 0.002 | 0.001 to 0.003 | <.001 |

95%CI: 95% Confidence Interval, ACEI: Angiotensin-Converting Enzyme Inhibitor, ARB: Angiotensin II Receptor Blocker.

### Appendix.
### List of the SEMI-COVID-19 Network members

**Coordinator of the SEMI-COVID-19 Registry:** José Manuel Casas Rojo.

**SEMI-COVID-19 Scientific Committee Members:** José Manuel Casas Rojo, José Manuel Ramos Rincón, Carlos Lumbreras Bermejo, Jesús Millán Núñez-Cortés, Juan Miguel Antón Santos, Ricardo Gómez Huelgas.

**SEMI-COVID-19 Registry Coordinating Center:** S & H Medical Science Service.

**APPENDIX**
Members of the SEMI-COVID-19 Group

H. U. 12 de Octubre. Madrid
Paloma Agudo de Blas, Coral Arévalo Cañas, Blanca Ayuso, José Bascuñana Morejón, Samara Campos Escudero, María Carnevali Frías, Santiago Cossio Tejido, Borja de Miguel Campo, Carmen Díaz Pedroche, Raquel Diaz Simon, Ana García Reyne, Lucia Jorge Huerta, Antonio Lalueza Blanco, Jaime Laureiro Gonzalo, Jaime Lora-Tamayo, Carlos Lumbreras Bermejo, Guillermo Maestro de la Calle, Barbara Otero Perpiña, Diana Paredes Ruiz, Marcos Sánchez Fernández, Javier Tejada Montes.

Hospital Universitari de Bellvitge. L'Hospitalet de Llobregat
Xavier Corbella, Narcís Homs, Abelardo Montero, Jose María Mora-Luján, Manuel Rubio-Rivas.

H. U. Gregorio Marañón. Madrid
Laura Abarca Casas, Álvaro Alejandre de Oña, Rubén Alonso Beato, Leyre Alonso Gonzalo, Jaime Alonso Muñoz, Crhistian Mario Amodeo Oblitas, Cristina Ausín García, Marta Bacete Cebrián, Jesús Baltasar Corral, Maria Barrientos Guerrero, Alejandro Bendala Estrada, María Calderón Moreno, Paula Carrascosa Fernández, Raquel Carrillo, Sabela Castañeda Pérez, Eva Cervilla Muñoz, Agustín Diego Chacón Moreno, Maria Carmen Cuenca Carvajal, Sergio de Santos, Andrés Enríquez Gómez, Eduardo Fernández Carracedo, María Mercedes Ferreiro-Mazón Jenaro, Francisco Galeano Valle, Alejandra Garcia, Irene Garcia Fernandez-Bravo, María Eugenia García Leoni, Maria Gomez Antunez, Candela González San Narciso, Anthony Alexander Gurjian, Lorena Jiménez Ibáñez, Cristina Lavilla Olleros, Cristina Llamazares Mendo, Sara Luis García, Víctor Mato Jimeno, Clara Millán Nohales, Jesús Millán Núñez-Cortés, Sergio Moragón Ledesma, Antonio Muiño Miguez, Cecilia Muñoz Delgado, Lucía Ordieres Ortega, Susana Pardo Sánchez, Alejandro Parra Virto, María Teresa Pérez Sanz, Blanca Pinilla Llorente, Sandra Piqueras Ruiz, Guillermo Soria Fernández-Llamazares, María Toledano Macías, Neera Toledo Samaniego, Ana Torres do Rego, Maria Victoria Villalba Garcia, Gracia Villarreal, María Zurita Etayo.

H. U. La Paz-Cantoblanco-Carlos III. Madrid

Jorge Álvarez Troncoso, Francisco Arnalich Fernández, Francisco Blanco Quintana, Carmen Busca Arenzana, Sergio Carrasco Molina, Aranzazu Castellano Candalija, Germán Daroca Bengoa, Alejandro de Gea Grela, Alicia de Lorenzo Hernández, Alejandro Díez Vidal, Carmen Fernández Capitán, Maria Francisca García Iglesias, Borja González Muñoz, Carmen Rosario Herrero Gil, Juan María Herrero Martínez, Víctor Hontañón, Maria Jesús Jaras Hernández, Carlos Lahoz, Cristina Marcelo Calvo, Juan Carlos Martín Gutiérrez, Monica Martinez Prieto, Elena Martínez Robles, Araceli Menéndez Saldaña, Alberto Moreno Fernández, Jose Maria Mostaza Prieto, Ana Noblejas Mozo, Carlos Manuel Oñoro López, Esmeralda Palmier Peláez, Marina Palomar Pampyn, Maria Angustias Quesada Simón, Juan Carlos Ramos Ramos, Luis Ramos Ruperto, Aquilino Sánchez Purificación, Teresa Sancho Bueso, Raquel Sorriguieta Torre, Clara Itziar Soto Abanedes, Yeray Untoria Tabares, Marta Varas Mayoral, Julia Vásquez Manau.

C. H. U. de Albacete. Albacete
Jose Luis Beato Pérez, Maria Lourdes Sáez Méndez.

H. U. Puerta de Hierro. Majadahonda
María Álvarez Bello, Ane Andrés Eisenhofer, Ana Arias Milla, Isolina Baños Pérez, Laura Benítez Gutiérrez, Javier Bilbao Garay, Silvia Blanco Alonso, Jorge Calderón Parra, Alejandro Callejas Díaz, José María Camino Salvador, Mª Cruz Carreño Hernández, Valentín Cuervas-Mons Martínez, Sara de la Fuente Moral, Miguel del Pino Jimenez, Alberto Díaz de Santiago, Itziar Diego Yagüe, Ignacio Donate Velasco, Ana María Duca, Pedro Durán del Campo, Gabriela Escudero López, Esther Expósito Palomo, Ana Fernández Cruz, Esther Fiz Benito, Andrea Fraile López, Amy Galán Gómez, Sonia García Prieto, Claudia García Rodríguez-Maimón, Miguel Ángel García Viejo, Javier Gómez Irusta, Edith Vanessa Gutiérrez Abreu, Isabel Gutiérrez Martín, Ángela Gutiérrez Rojas, Andrea Gutiérrez Villanueva, Jesús Herráiz Jiménez, Pedro Laguna del Estal, Mª Carmen Máinez Sáiz, Cristina Martín Martín, María Martínez Urbistondo, Fernando Martínez Vera, Susana Mellor Pita, Patricia Mills Sánchez, Esther Montero Hernández, Alberto Mora Vargas, Cristina Moreno López, Alfonso Ángel-Moreno Maroto, Victor Moreno-Torres Concha, Ignacio Morrás De La Torre, Elena Múñez Rubio, Ana Muñoz Gómez, Rosa Muñoz de Benito, Alejandro Muñoz Serrano, Jose María Palau Fayós, Lina Marcela Parra Ramírez, Ilduara Pintos Pascual, Arturo José Ramos Martín-Vegue, Antonio Ramos Martínez, Isabel Redondo Cánovas del Castillo, Alberto Roldán Montaud, Lucía Romero Imaz, Yolanda Romero Pizarro, Mónica Sánchez Santiuste, David Sánchez Órtiz, Enrique Sánchez Chica, Patricia Serrano de la Fuente, Pablo Tutor de Ureta, Ángela Valencia Alijo, Mercedes Valentín-Pastrana Aguilar, Juan Antonio Vargas Núñez, Jose Manuel Vázquez Comendador, Gema Vázquez Contreras, Carmen Vizoso Gálvez.

H. Miguel Servet. Zaragoza
Gonzalo Acebes Repiso, Uxua Asín Samper, María Aranzazu Caudevilla Martínez, José Miguel García Bruñén, Rosa García Fenoll, Jesús Javier González Igual, Laura Letona Giménez, Mónica Llorente Barrio.

Hospital Royo Villanova. Zaragoza
Nicolás Alcalá Rivera, Anxela Crestelo Vieitez, Esther del Corral Beamonte, Jesús Díez Manglano, Isabel Fiteni Mera, Maria del Mar Garcia Andreu, Martin Gerico Aseguinolaza, Claudia Josa Laorden, Raul Martínez Murgui, Marta Teresa Matía Sanz.

H. Clínico San Carlos. Madrid

Inés Armenteros Yeguas, Javier Azaña Gómez, Julia Barrado Cuchillo, Irene Burruezo López, Noemí Cabello Clotet, Alberto E. Calvo Elías, Elpidio Calvo Manuel, Verónica Cano, Carmen María Cano de Luque, Cynthia Chocron Benbunan, Laura Dans Vilan, Ester Emilia Dubon Peralta, Vicente Estrada Pérez, Santiago Fernandez-Castelao, Marcos Oliver Fragiel Saavedra, José Luis García Klepzig, Maria del Rosario Iguarán Bermúdez, Esther Jaén Ferrer, Alejandro Maceín Rodríguez, Rubén Ángel Martín Sánchez, Manuel Méndez Bailón, Sara Miguel Álvarez, Maria José Nuñez Orantos, Carolina Olmos Mata, Eva Orviz García, David Oteo Mata, Cristina Outon González, Juncal Perez-Somarriba, Pablo Pérez Mateos, Maria Esther Ramos Muñoz, Xabier Rivas Regaira, Laura Mª Rodríguez Gallardo, Iñigo Sagastagoitia Fornie, Alejandro Salinas Botrán, Miguel Suárez Robles, Maddalena Elena Urbano, Miguel Villar Martínez.

H. U. La Princesa. Madrid

María Aguilera García, Ester Alonso Monge, Jesús Álvarez Rodríguez, Claudia Alvarez Varela, Miquel Berniz Gòdia, Marta Briega Molina, Marta Bustamante Vega, Jose Curbelo, Alicia de las Heras Moreno, Ignacio Descalzo Godoy, Alexia Constanza Espiño Alvarez, Ignacio Fernández Martín-Caro, Alejandra Franquet López-Mosteiro, Gonzalo Galvez Marquez, María J. García Blanco, Yaiza García del Álamo Hernández, Clara García-Rayo Encina, Noemí Gilabert González, Carolina Guillamo Rodríguez, Nicolás Labrador San Martín, Manuel Molina Báez, Carmen Muñoz Delgado, Pedro Parra Caballero, Javier Pérez Serrano, Laura Rabes Rodríguez, Pablo Rodríguez Cortés, Carlos Rodriguez Franco, Emilia Roy-Vallejo, Monica Rueda Vega, Aresio Sancha Lloret, Beatriz Sánchez Moreno, Marta Sanz Alba, Jorge Serrano Ballester, Alba Somovilla, Carmen Suarez Fernández, Macarena Vargas Tirado, Almudena Villa Marti.

H. U. de A Coruña. A Coruña

Alicia Alonso Álvarez, Olaya Alonso Juarros, Ariadna Arévalo López, Carmen Casariego Castiñeira, Ana Cerezales Calviño, Marta Contreras Sánchez, Ramón Fernández Varela, Santiago J. Freire Castro, Ana Padín Trigo, Rafael Prieto Jarel, Fátima Raad Varea, Ignacio Ramil Freán, Laura Ramos Alonso, Francisco Javier Sanmartín Pensado, David Vieito Porto.

H. Moisès Broggi. Sant Joan Despí

Judit Aranda Lobo, Jose Loureiro Amigo, Isabel Oriol Bermúdez, Melani Pestaña Fernández, Nicolas Rhyman, Nuria Vázquez Piqueras.

Hospital Clínico de Santiago. Santiago de Compostela

Maria del Carmen Beceiro Abad, Maria Aurora Freire Romero, Sonia Molinos Castro, Emilio Manuel Paez Guillan, María Pazo Nuñez, Paula Maria Pesqueira Fontan.

Hospital Universitario Dr. Peset. Valencia

Juan Alberto Aguilera Ayllón, Arturo Artero, María del Mar Carmona Martín, María José Fabiá Valls, Maria de Mar Fernández Garcés, Ana Belén Gómez Belda, Ian López Cruz, Manuel Madrazo López, Elisabeth Mateo Sanchis, Jaume Micó Gandia, Laura Piles Roger, Adela Maria Pina Belmonte, Alba Viana García.

H. de Cabueñes. Gijón

Ana María Álvarez Suárez, Carlos Delgado Vergés, Rosa Fernandez-Madera Martínez, Eva Fonseca Aizpuru, Alejandro Gómez Carrasco, Cristina Helguera Amezua, Juan Francisco López Caleya, María del Mar Martínez López, Aleida Martínez Zapico, Carmen Olabuenaga Iscar, María Luisa Taboada Martínez, Lara María Tamargo Chamorro.

H. U. Ramón y Cajal. Madrid
Luis FernandoAbrego Vaca, Ana Andréu Arnanz, Octavio Arce García, Marta Bajo González, Pablo Borque Sanz, Alberto Cozar Llisto, Sonia de Pedro Baena, Beatriz Del Hoyo Cuenda, María Alejandra Gamboa Osorio, Isabel García Sánchez, Andrés González García, Oscar Alberto López Cisneros, Miguel Martínez Lacalzada, Borja Merino Ortiz, Jimena Rey-García, Elisa Riera González, Cristina Sánchez Díaz, Grisell Starita Fajardo, Cecilia Suárez Carantoña, Adrian Viteri Noel, Svetlana Zhilina Zhilina.

H. Nuestra Señora del Prado. Talavera de la Reina
Sonia Casallo Blanco, Jeffrey Oskar Magallanes Gamboa.

C. Asistencial de Zamora. Zamora
Carlos Aldasoro Frias, Luis Arribas Perez, María Esther Fraile Villarejo, Beatriz Garcia Lopez, Victor Madrid Romero, Emilia Martínez Velado, Victoria Palomar Calvo, Sara Pintos Otero, Carlota Tuñón de Almeida

H. Virgen de la Salud. Toledo
Ana Maria Alguacil Muñoz, Marta Blanco Fernández, Veronica Cano, Ricardo Crespo Moreno, Fernando Cuadra Garcia-Tenorio, Blanca Díaz-Tendero Nájera, Raquel Estévez González, María Paz García Butenegro, Alberto Gato Díez, Verónica Gómez Caverzaschi, Piedad María Gómez Pedraza, Julio González Moraleja, Raúl Hidalgo Carvajal, Patricia Jiménez Aranda, Raquel Labra González, Áxel Legua Caparachini, Pilar Lopez Castañeyra, Agustín Lozano Ancin, Jose Domingo Martin Garcia, Cristina Morata Romero, María Jesús Moya Saiz, Helena Moza Moríñigo, Gemma Muñiz Nicolás, Enriqueta Muñoz Platon, Filomena Oliveri, Elena Ortiz Ortiz, Raúl Perea Rafael, Pilar Redondo Galán, María Antonia Sepulveda Berrocal, Vicente Serrano Romero de Ávila, Pilar Toledano Sierra, Yamilex Urbano Aranda, Jesús Vázquez Clemente, Carmen Yera Bergua.

H. U. Infanta Cristina. Parla
Juan Miguel Antón Santos, Ana Belén Barbero Barrera, Coralia Bueno Muiño, Ruth Calderón Hernaiz, Irene Casado Lopez, José Manuel Casas Rojo, Andrés Cortés Troncoso, Mayte de Guzmán García-Monge, Francesco Deodati, Gonzalo García Casasola Sánchez, Elena Garcia Guijarro, Davide Luordo, María Mateos González, Jose A Melero Bermejo, Lorea Roteta García, Elena Sierra Gonzalo, Javier Villanueva Martínez.

Hospital Regional Universitario de Málaga. Málaga
Mª Mar Ayala Gutiérrez, Rosa Bernal López, José Bueno Fonseca, Verónica Andrea Buonaiuto, Luis Francisco Caballero Martínez, Lidia Cobos Palacios, Clara Costo Muriel, Francis de Windt, Ana Teresa Fernandez-Truchaud Christophel, Paula García Ocaña, Ricardo Gómez Huelgas, Javier Gorospe García, Maria Dolores López Carmona, Pablo López Quirantes, Almudena López Sampalo, Elizabeth Lorenzo Hernández, Juan José Mancebo Sevilla, Jesica Martin Carmona, Luis Miguel Pérez-Belmonte, Araceli Pineda Cantero, Carlos Romero Gómez, Michele Ricci, Jaime Sanz Cánovas

H. U. San Juan de Alicante. San Juan de Alicante
Marisa Asensio Tomás, David Balaz, David Bonet Tur, Ruth Cañizares Navarro, Paloma Chazarra Pérez, Jesús Corbacho Redondo, Leticia Espinosa Del Barrio, Pedro Jesús Esteve Atiénzar, Carles García Cervera, David Francisco García Núñez, Vicente Giner Galvañ, Angie Gómez Uranga, Javier Guzmán Martínez, Isidro Hernández Isasi, Lourdes Lajara Villar, Verónica Martínez Sempere, Juan Manuel Núñez Cruz, Sergio Palacios Fernández, Juan Jorge Peris García, Andrea Riaño Pérez, José Miguel Seguí Ripoll, Azucena Sempere Mira, Philip Wikman-Jorgensen.

Hospital Costa del Sol. Marbella
Nicolás Jiménez García, Jairo Luque del Pino, María Dolores Martín Escalante.

H. del Henares. Coslada
Jesús Ballano Rodríguez-Solís, Luis Cabeza Osorio, María del Pilar Fidalgo Montero, Mª Isabel Fuentes Soriano, Erika Esperanza Lozano Rincon, Ana Martín Hermida, Jesus Martinez Carrilero, Jose Angel Pestaña Santiago, Manuel Sánchez Robledo, Patricia Sanz Rojas, Nahum Jacobo Torres Yebes, Vanessa Vento.

H. U. La Fe. Valencia
Dafne Cabañero, María Calabuig Ballester, Pascual Císcar Fernández, Ricardo Gil Sánchez, Marta Jiménez Escrig, Cristina Marín Amela, Laura Parra Gómez, Carlos Puig Navarro, José Antonio Todolí Parra.

H. de Mataró. Mataró
Raquel Aranega González, Ramon Boixeda, Javier Fernández Fernández, Carlos Lopera Mármol, Marta Parra Navarro, Ainhoa Rex Guzmán, Aleix Serrallonga Fustier.

H. San Pedro. Logroño
Diana Alegre González, Irene Ariño Pérez de Zabalza, Sergio Arnedo Hernández, Jorge Collado Sáenz, Beatriz Dendariena, Marta Gómez del Mazo, Iratxe Martínez de Narvajas Urra, Sara Martínez Hernández, Estela Menendez Fernández, Jose Luís Peña Somovilla, Elisa Rabadán Pejenaute.

Complejo Hospitalario Universitario Ourense. Ourense
Raquel Fernández González, Amara Gonzalez Noya, Carlos Hernández Ceron, Isabel Izuzquiza Avanzini, Ana Latorre Diez, Pablo López Mato, Ana María Lorenzo Vizcaya, Daniel Peña Benítez, Milagros María Peña Zemsch, Lucía Pérez Expósito, Marta Pose Bar, Lara Rey González, Laura Rodrigo Lara

H. U. Reina Sofía. Córdoba
Antonio Pablo Arenas de Larriva, Pilar Calero Espinal, Javier Delgado Lista, Francisco Fuentes-Jiménez, María Jesús Gómez Vázquez, Jose Jiménez Torres, José López-Miranda, Laura Martín Piedra, Javier Pascual Vinagre, Pablo Pérez-Martinez, María Elena Revelles Vílchez, Juan Luis Romero Cabrera, José David Torres Peña.

H. Juan Ramón Jiménez. Huelva

Francisco Javier Bejarano Luque, Francisco Javier Carrasco-Sánchez, Mercedes de Sousa Baena, Jaime Díaz Leal, Aurora Espinar Rubio, Maria Franco Huertas, Juan Antonio García Bravo, Andrés Gonzalez Macías, Encarnación Gutiérrez Jiménez, Alicia Hidalgo Jiménez, Constantino Lozano Quintero, Carmen Mancilla Reguera, Francisco Javier Martínez Marcos, Francisco Muñoz Beamud, Maria Perez Aguilera, Alícia Perez Jiménez, Virginia Rodríguez Castaño, Alvaro Sánchez de Alcazar del Río, Leire Toscano Ruiz.

Hospital Alto Guadalquivir. Andújar
Begoña Cortés Rodríguez.

Hospital Infanta Margarita. Cabra
María Esther Guisado Espartero, Lorena Montero Rivas, Maria de la Sierra Navas Alcántara, Raimundo Tirado-Miranda.

C. H. U. de Ferrol. Ferrol
Hortensia Alvarez Diaz, Tamara Dalama Lopez, Estefania Martul Pego, Carmen Mella Pérez, Ana Pazos Ferro, Sabela Sánchez Trigo, Dolores Suarez Sambade, Maria Trigas Ferrin, Maria del Carmen Vázquez Friol, Laura Vilariño Maneiro.

Complejo Asistencial Universitario de León. León
Rosario Maria García Diez, Manuel Martin Regidor, Angel Luis Martínez Gonzalez, Alberto Muela Molinero, Raquel Rodríguez Díez, Beatriz Vicente Montes.

Hospital Marina Baixa. Villajoyosa
Javier Ena, Jose Enrique Gómez Segado.

Hospital Torrecárdenas. Almería
Luis Felipe Díez García, Iris El Attar Acedo, Bárbara Hernandez Sierra, Carmen Mar Sánchez Cano.

H. U. Severo Ochoa. Leganés
Yolanda Casillas Viera, Lucía Cayuela Rodríguez, Carmen de Juan Alvarez, Gema Flox Benitez, Laura García Escudero, Juan Martin Torres, Patricia Moreira Escriche, Susana Plaza Canteli, M Carmen Romero Pérez.

C. H. U. de Badajoz. Badajoz
Rafael Aragon Lara, Inmaculada Cimadevilla Fernandez, Juan Carlos Cira García, Gema Maria García García, Julia Gonzalez Granados, Beatriz Guerrero Sánchez, Francisco Javier Monreal Periáñez, Maria Josefa Pascual Perez.

H. de Pozoblanco. Pozoblanco
José Nicolás Alcalá Pedrajas, Antonia Márquez García, Inés Vargas.

Hospital Platón. Barcelona
Ana Suarez Lombraña

Hospital Valle del Nalón. Riaño (Langreo)
Sara Fuente Cosío, César Manuel Gallo Álvaro, Julia Lobo García, Antía Pérez Piñeiro.

H. General Defensa
Anyuli Gracia Gutiérrez, Leticia Esther Royo Trallero

H. U. del Vinalopó. Elche
Francisco Amorós Martínez, Erika Ascuña Vásquez, José Carlos Escribano Stablé, Adriana Hernández Belmonte, Ana Maestre Peiró, Raquel Martínez Goñi, M.Carmen Pacheco Castellanos, Bernardino Soldan Belda, David Vicente Navarro.

H. Francesc de Borja. Gandia
Alba Camarena Molina, Simona Cioaia, Anna Ferrer Santolalia, José María Frutos Pérez, Eva Gil Tomás, Leyre Jorquer Vidal, Marina Llopis Sanchis, M Ángeles Martínez Pascual, Alvaro Navarro Batet, Mari Amparo Perea Ribis, Ricardo Peris Sanchez, José Manuel Querol Ribelles, Silvia Rodriguez Mercadal, Ana Ventura Esteve.

H. G. U. de Castellón. Castellón de la Plana
Jorge Andrés Soler, Marián Bennasar Remolar, Alejandro Cardenal Álvarez, Daniela Díaz Carlotti, María José Esteve Gimeno, Sergio Fabra Juana, Paula García López, María Teresa Guinot Soler, Daniela Palomo de la Sota, Guillem Pascual Castellanos, Ignacio Pérez Catalán, Celia Roig Martí, Paula Rubert Monzó, Javier Ruiz Padilla, Nuria Tornador Gaya, Jorge Usó Blasco.

C. A. U. de Salamanca. Salamanca
Gloria María Alonso Claudio, Víctor Barreales Rodríguez, Cristina Carbonell Muñoz, Adela Carpio Pérez, María Victoria Coral Orbes, Daniel Encinas Sánchez, Sandra Inés Revuelta, Miguel Marcos Martín, José Ignacio Martín González, José Ángel Martín Oterino, Leticia Moralejo Alonso, Sonia Peña Balbuena, María Luisa Pérez García, Ana Ramon Prados, Beatriz Rodríguez-Alonso, Ángela Romero Alegría, Maria Sanchez Ledesma, Rosa Juana Tejera Pérez.

H. U. Quironsalud Madrid. Pozuelo de Alarcón (Madrid)
Pablo Guisado Vasco, Ana Roda Santacruz, Ana Valverde Muñoz.

H. U. del Sureste. Arganda del Rey
Jon Cabrejas Ugartondo, Ana Belén Mancebo Plaza, Arturo Noguerado Asensio, Bethania Pérez Alves, Natalia Vicente López.

Hospital de Palamós. Palamós
Ana Alberich Conesa, Maricruz Almendros Rivas, Miquel Hortos Alsina, Jorge Marchena Romero, Anabel Martin-Urda Diez-Canseco.

H. Parc Tauli. Sabadell
Francisco Epelde, Isabel Torrente

Hospital Doctor José Molina Orosa. Arrecife (Lanzarote)
Virginia Herrero García, Berta Román Bernal.

H. Virgen de los Lirios. Alcoy (Alicante)

Mª José Esteban Giner.

Hospital Clínico Universitario de Valladolid. Valladolid
Xjoylin Teresita Egües Torres, Sara Gutiérrez González, Cristina Novoa Fernández, Pablo Tellería Gómez.

Hospital Público de Monforte de Lemos. Monforte de Lemos
José López Castro, Manuel Lorenzo López Reboiro

H. Virgen del Mar. Madrid
Thamar Capel Astrua, Paola Tatiana Garcia Giraldo, Maria Jesús González Juárez, Victoria Marquez Fernandez, Ada Viviana Romero Echevarry.

Hospital do Salnes. Vilagarcía de Arousa
Vanesa Alende Castro, Ana María Baz Lomba, Ruth Brea Aparicio, Marta Fernandez Morales, Jesús Manuel Fernández Villar, María Teresa López Monteagudo, Cristina Pérez García, Lorena Rodríguez Ferreira, Diana Sande Llovo, Maria Begoña Valle Feijoo.

**STROBE (Strengthening The Reporting of OBservational Studies in Epidemiology) Checklist**

A checklist of items that should be included in reports of observational studies. You must report the page number in your manuscript where you consider each of the items listed in this checklist. If you have not included this information, either revise your manuscript accordingly before submitting or note N/A.

**Note:** An Explanation and Elaboration article discusses each checklist item and gives methodological background and published examples of transparent reporting. The STROBE checklist is best used in conjunction with this article (freely available on the Web sites of PLoS Medicine at http://www.plosmedicine.org/, Annals of Internal Medicine at http://www.annals.org/, and Epidemiology at http://www.epidem.com/). Information on the STROBE Initiative is available at www.strobe-statement.org.

| Section and Item | Item No. | Recommendation | Reported on Page No. |
|---|---|---|---|
| **Title and Abstract** | 1 | (*a*) Indicate the study's design with a commonly used term in the title or the abstract | 1 |
| | | (*b*) Provide in the abstract an informative and balanced summary of what was done and what was found | 4 |
| **Introduction** | | | |
| Background/Rationale | 2 | Explain the scientific background and rationale for the investigation being reported | 5 |
| Objectives | 3 | State specific objectives, including any prespecified hypotheses | 5 |
| **Methods** | | | |
| Study Design | 4 | Present key elements of study design early in the paper | 6 |
| Setting | 5 | Describe the setting, locations, and relevant dates, including periods of recruitment, exposure, follow-up, and data collection | 6 |
| Participants | 6 | (*a*) *Cohort study*—Give the eligibility criteria, and the sources and methods of selection of participants. Describe methods of follow-up | 6 |
| | | *Case-control study*—Give the eligibility criteria, and the sources and methods of case ascertainment and control selection. Give the rationale for the choice of cases and controls | NA |
| | | *Cross-sectional study*—Give the eligibility criteria, and the sources and methods of selection of participants | NA |
| | | (*b*) *Cohort study*—For matched studies, give matching criteria and number of exposed and unexposed | 7-9 |
| | | *Case-control study*—For matched studies, give matching criteria and the number of controls per case | NA |
| Variables | 7 | Clearly define all outcomes, exposures, predictors, potential confounders, and effect modifiers. Give diagnostic criteria, if applicable | 7 |

| Section and Item | Item No. | Recommendation | Reported on Page No. |
|---|---|---|---|
| Data Sources/ Measurement | 8* | For each variable of interest, give sources of data and details of methods of assessment (measurement). Describe comparability of assessment methods if there is more than one group | 7 |
| Bias | 9 | Describe any efforts to address potential sources of bias | 7,8 |
| Study Size | 10 | Explain how the study size was arrived at | 6, Figure 1 |
| Quantitative Variables | 11 | Explain how quantitative variables were handled in the analyses. If applicable, describe which groupings were chosen and why | 7,8 |
| Statistical Methods | 12 | (*a*) Describe all statistical methods, including those used to control for confounding | 7,8 |
| | | (*b*) Describe any methods used to examine subgroups and interactions | 7,8 |
| | | (*c*) Explain how missing data were addressed | 7,8 |
| | | (*d*) *Cohort study*—If applicable, explain how loss to follow-up was addressed | 6 |
| | | *Case-control study*—If applicable, explain how matching of cases and controls was addressed | NA |
| | | *Cross-sectional study*—If applicable, describe analytical methods taking account of sampling strategy | NA |
| | | (*e*) Describe any sensitivity analyses | 7,8 |
| **Results** | | | |
| Participants | 13* | (a) Report numbers of individuals at each stage of study—eg numbers potentially eligible, examined for eligibility, confirmed eligible, included in the study, completing follow-up, and analysed | 9, Figure 1 |
| | | (b) Give reasons for non-participation at each stage | 9, Figure 1 |
| | | (c) Consider use of a flow diagram | 9,Figure 1 |
| Descriptive Data | 14* | (a) Give characteristics of study participants (eg demographic, clinical, social) and information on exposures and potential confounders | 9, Table 1 |
| | | (b) Indicate number of participants with missing data for each variable of interest | Table 1 |
| | | (c) *Cohort study*—Summarise follow-up time (eg, average and total amount) | 6 |
| Outcome Data | 15* | *Cohort study*—Report numbers of outcome events or summary measures over time | 9-11, Tables 2 |
| | | *Case-control study*—Report numbers in each exposure category, or summary measures of exposure | NA |
| | | *Cross-sectional study*—Report numbers of outcome events or summary measures | NA |

| Section and Item | Item No. | Recommendation | Reported on Page No. |
|---|---|---|---|
| Main Results | 16 | (*a*) Give unadjusted estimates and, if applicable, confounder-adjusted estimates and their precision (eg, 95% confidence interval). Make clear which confounders were adjusted for and why they were included | 9-11 |
| | | (*b*) Report category boundaries when continuous variables were categorized | NA |
| | | (*c*) If relevant, consider translating estimates of relative risk into absolute risk for a meaningful time period | NA |
| Other Analyses | 17 | Report other analyses done—eg analyses of subgroups and interactions, and sensitivity analyses | 9-11 |
| **Discussion** | | | |
| Key Results | 18 | Summarise key results with reference to study objectives | 11-14 |
| Limitations | 19 | Discuss limitations of the study, taking into account sources of potential bias or imprecision. Discuss both direction and magnitude of any potential bias | 14,15 |
| Interpretation | 20 | Give a cautious overall interpretation of results considering objectives, limitations, multiplicity of analyses, results from similar studies, and other relevant evidence | 15 |
| Generalisability | 21 | Discuss the generalisability (external validity) of the study results | 15 |
| **Other Information** | | | |
| Funding | 22 | Give the source of funding and the role of the funders for the present study and, if applicable, for the original study on which the present article is based | 15 |

*Give information separately for cases and controls in case-control studies and, if applicable, for exposed and unexposed groups in cohort and cross-sectional studies.

**Once you have completed this checklist, please save a copy and upload it as part of your submission. DO NOT include this checklist as part of the main manuscript document. It must be uploaded as a separate file.**
